# Evaluating Scenarios for School Reopening under COVID19

**DOI:** 10.1101/2020.07.22.20160036

**Authors:** Arden Baxter, Buse Eylul Oruc, Pinar Keskinocak, John Asplund, Nicoleta Serban

## Abstract

Thousands of school systems have been struggling with the decisions about how to safely and effectively deliver education during the fall semester of 2020, amid the COVID19 pandemic. The objective of this study is to evaluate the public health impact of reopening schools on the spread of COVID19. An agent-based simulation model was adapted and used to project the number of infections and deaths under multiple school reopening dates and scenarios, including different cohorts receiving in-person instruction on alternating days, only younger children returning to in-person instruction, regular schedule (all students receiving in-person instruction), and school closure (all students receiving online instruction). The study period was February 18^th^-November 24^th^, 2020 and the state of Georgia was used as a case study. Across all scenarios, the number of COVID19-related deaths ranged from approximately 17 to 22 thousand during the study period, and on the peak day, the number of new infections ranged from 43 to 68 thousand. An alternating school day schedule performed: (i) almost as well as keeping schools closed, with the infection attack rate ranging from 38.5% to 39.8% compared to that of 37.7% under school closure; (ii) slightly better than only allowing children 10 years or younger to return to in-person instruction. Delaying the reopening of schools had a minimal impact on reducing infections and deaths under most scenarios.

**Significance Statement:** This study provides insights on the impact of various school reopening dates and scenarios on the spread of COVID19, incorporating differences between children and adults in terms of disease progression and community transmission. School districts are faced with these challenging decisions considering the complex tradeoffs of their impact between public health, education, and society. While the number of new COVID19 confirmed cases continue to increase in many states, so are concerns about the negative impact of school closures on the children’s education and development. The systematic analysis of school reopening scenarios provided in this study will support school systems in their decision-making regarding if, when, and how to return to in-person instruction.

## Introduction

School systems have been developing plans for safely reopening during the fall semester of 2020 while considering the potential impact of in-person interactions on students, staff, families, and public health during the COVID19 pandemic (1–3). Recent studies have shown the potential benefits of non-pharmaceutical interventions, such as school closures (4, 5), in slowing down infection spread and reducing the severe health outcomes, but also highlighted their negative impact on the economy, unemployment, mobility, mental health, education, caregiving, etc. (6, 7). Widespread school closures during spring 2020 not only impacted the education of children and youth but also had economic consequences due to increased childcare responsibilities of working parents (8–16).

To evaluate the tradeoffs between the potential public health benefits of various school reopening scenarios versus their impact on the educational development of children and the economy, in this study, we considered intervention metrics, namely, the projected number of infections and deaths due to COVID19, and the following school reopening scenarios during the fall semester of 2020: (a) *schools closed* schedule: all students receive online instruction; (b) *alternating school day for younger children* schedule: only children 10 years old or younger return to in-person instruction while following an alternating school day schedule; (c) *alternating school day* schedule: half of the students receive in-person instruction on Mondays and Wednesdays and the other half on Tuesdays and Thursdays; (d) *younger children only* schedule: only children 10 years old or younger return to in-person instruction; (e) *regular* schedule: all students return to in-person instruction.

Quantifying the public health benefits of school reopening scenarios aim to provide much needed insights for school system decision-makers.

## Results

All the results presented utilized data from the state of Georgia, which has a total population of approximately 10.8 million where 1.3 million children are of age 0-9 and 1.4 million children are of age 10-19 (17). Kindergarten through 12^th^ grade (K-12) schools typically open during the first or second week of August in Georgia; hence, the following reopening dates were considered: August 10, August 17, August 24, August 31, September 7, and September 14. *Figure 1* depicts the base scenario.

**Figure 1.**
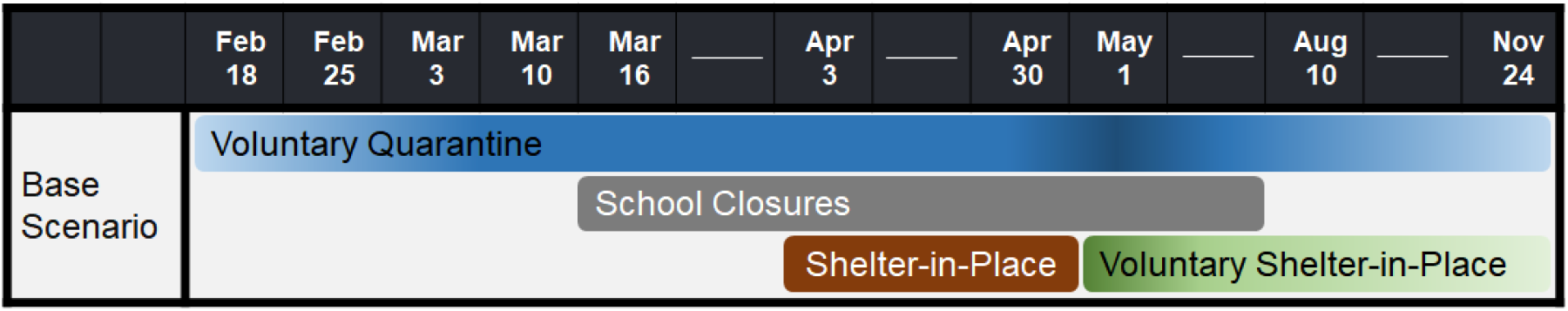
All scenarios considered are built upon the base scenario along with the corresponding school reopening date. Compliance with shelter-in-place, voluntary quarantine, and voluntary shelter-in-place varies over time.

Depending on the school reopening date, scenario, and the public’s participation in physical distancing, the number of COVID19-related deaths by November 24 could range from approximately 17.4 to 22 thousand in the state of Georgia. On the peak day (when the number of new infections on a given day is highest), the number of daily new infections could range from 19.4 to 47.6 thousand for adults and 10.6 to 22.2 thousand for children.

For the reopening date of August 10, in scenarios (a) through (e), respectively:

- The cumulative number of infections was (approximately, in thousands) 3,0377; 3,098; 3,166; 3,242; 3,600 for adults and 1,037; 1,072; 1,134; 1,183; 1,491 for children.
- The cumulative number of deaths was 17,417; 18,075; 18,385; 18,977; 21,980.
- The peak number of infections was 43,360; 45,466; 46,609; 48,836; 67,896.
- The peak day was August 19; August 18; August 23; August 24; August 30.

Results for all other metrics (IAR %, i.e., the percentage of the population infected, peak infections for adults and children and peak day) for other reopening dates can be found in *Table 1*. The relative ordering of the scenarios remains as (a) through (e) for each reopening date, with (a) being lowest and (e) being highest regarding infections and deaths. *Figures 2* and *3* present a comparison of the daily number of new infections under different school reopening scenarios.

**Table 1.** Summary comparison of school reopening scenarios with respect to cumulative deaths, cumulative infections in adults and children, infection attack rate (IAR %), peak infections in adults and children, and peak day.

| Schedules | Cumulative Deaths | Cumulative Infections |  | IAR % | Peak Infections |  | Peak Day |
| --- | --- | --- | --- | --- | --- | --- | --- |
|  |  | Adult | Children |  | Adults | Children |  |
| **Schools Closed** | 17417 | 3036819 | 1037471 | 37.73 | 32562 | 10798 | 19-Aug |
| **Regular 1** | 21980 | 3600338 | 1490883 | 47.15 | 45750 | 22146 | 30-Aug |
| **Regular 2** | 21405 | 3566364 | 1455359 | 46.51 | 40718 | 20099 | 2-Sep |
| **Regular 3** | 20902 | 3516434 | 1412750 | 45.65 | 35376 | 17845 | 8-Sep |
| **Regular 4** | 20318 | 3451780 | 1361897 | 44.58 | 29626 | 15141 | 10-Sep |
| **Regular 5** | 19791 | 3375957 | 1302854 | 43.33 | 32834 | 10834 | 15-Aug |
| **Regular 6** | 19346 | 3320594 | 1259850 | 42.42 | 32647 | 10774 | 18-Aug |
| **Alternating School Day 1** | 18385 | 3165649 | 1133661 | 39.82 | 34229 | 12380 | 23-Aug |
| **Alternating School Day 2** | 18466 | 3164226 | 1121944 | 39.69 | 33241 | 12159 | 22-Aug |
| **Alternating School Day 3** | 18264 | 3154783 | 1116358 | 39.55 | 32000 | 14348 | 25-Aug |
| **Alternating School Day 4** | 17912 | 3111939 | 1094476 | 38.95 | 32717 | 10651 | 19-Aug |
| **Alternating School Day 5** | 18030 | 3115832 | 1089622 | 38.95 | 33427 | 11081 | 18-Aug |
| **Alternating School Day 6** | 17739 | 3086135 | 1072150 | 38.51 | 32717 | 10769 | 19-Aug |
| **Younger Children Only 1** | 18977 | 3242205 | 1183429 | 40.99 | 35232 | 13604 | 24-Aug |
| **Younger Children Only 2** | 18813 | 3228035 | 1168330 | 40.71 | 33609 | 12958 | 25-Aug |
| **Younger Children Only 3** | 18652 | 3195697 | 1148196 | 40.23 | 32714 | 10663 | 18-Aug |
| **Younger Children Only 4** | 18499 | 3164906 | 1129067 | 39.77 | 32659 | 10799 | 19-Aug |
| **Younger Children Only 5** | 18333 | 3154983 | 1115030 | 39.54 | 33395 | 11048 | 18-Aug |
| **Younger Children Only 6** | 18030 | 3126636 | 1097190 | 39.12 | 33036 | 10815 | 19-Aug |
| **Alternating School Day for Younger Children 1** | 18075 | 3098238 | 1072097 | 38.62 | 33939 | 11527 | 18-Aug |
| **Alternating School Day for Younger Children 2** | 17852 | 3081321 | 1062270 | 38.37 | 32608 | 11097 | 20-Aug |
| **Alternating School Day for Younger Children 3** | 17707 | 3076859 | 1060334 | 38.31 | 32832 | 10759 | 17-Aug |
| **Alternating**<br>**School Day for**<br>**Younger Children**<br>**4** | 17866 | 3078539 | 1057583 | 38.30 | 33093 | 10831 | 17-Aug |
| **Alternating**<br>**School Day for**<br>**Younger Children**<br>**5** | 17736 | 3071110 | 1055003 | 38.21 | 33501 | 11018 | 17-Aug |
| **Alternating**<br>**School Day for**<br>**Younger Children**<br>**6** | 17508 | 3057110 | 1048999 | 38.03 | 32513 | 10724 | 16-Aug |

**Figure 2.**
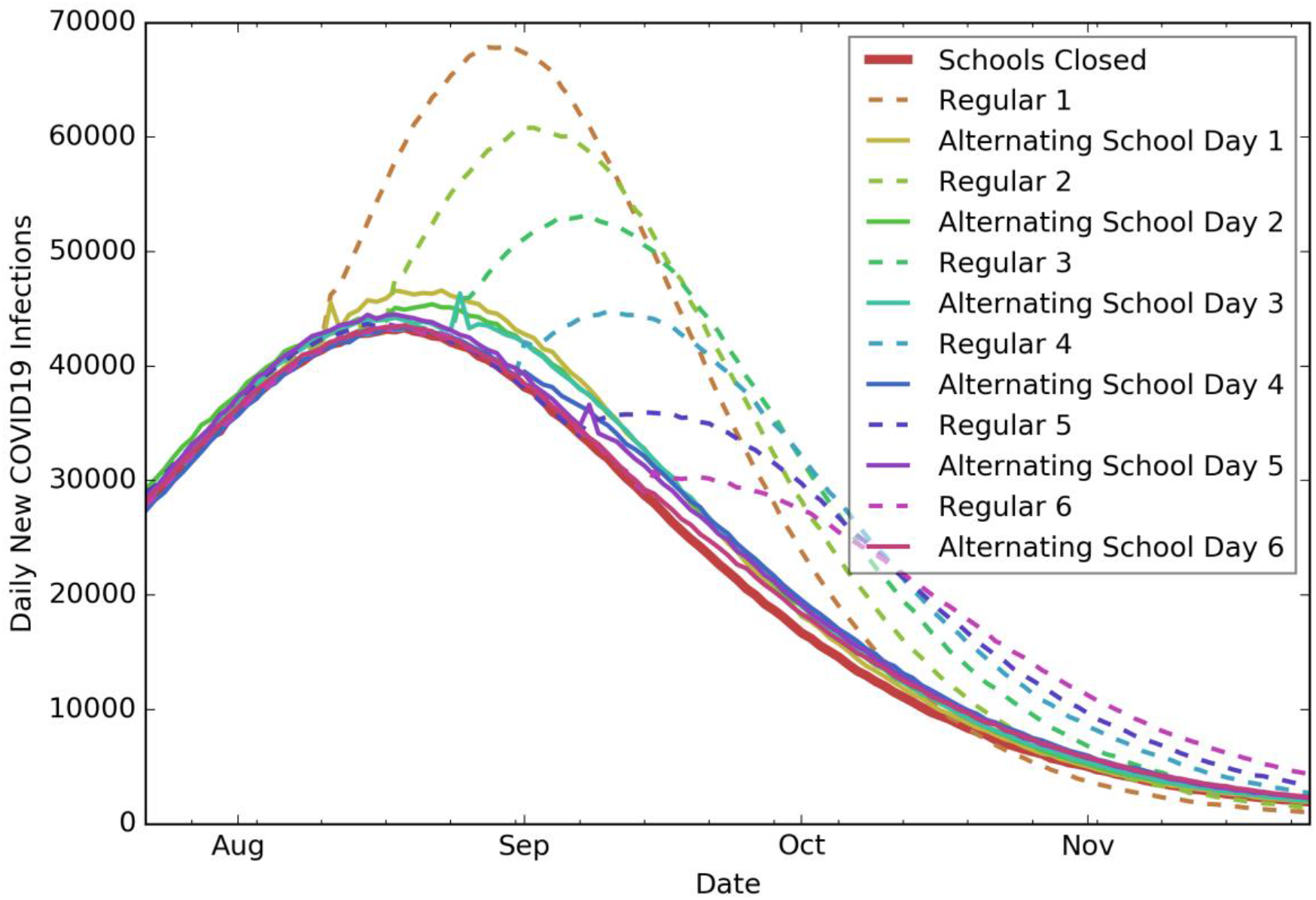
Daily new COVID19 infections under the *schools closed* schedule, the *regular* schedule, and the *alternating school day* schedule.

**Figure 3.**
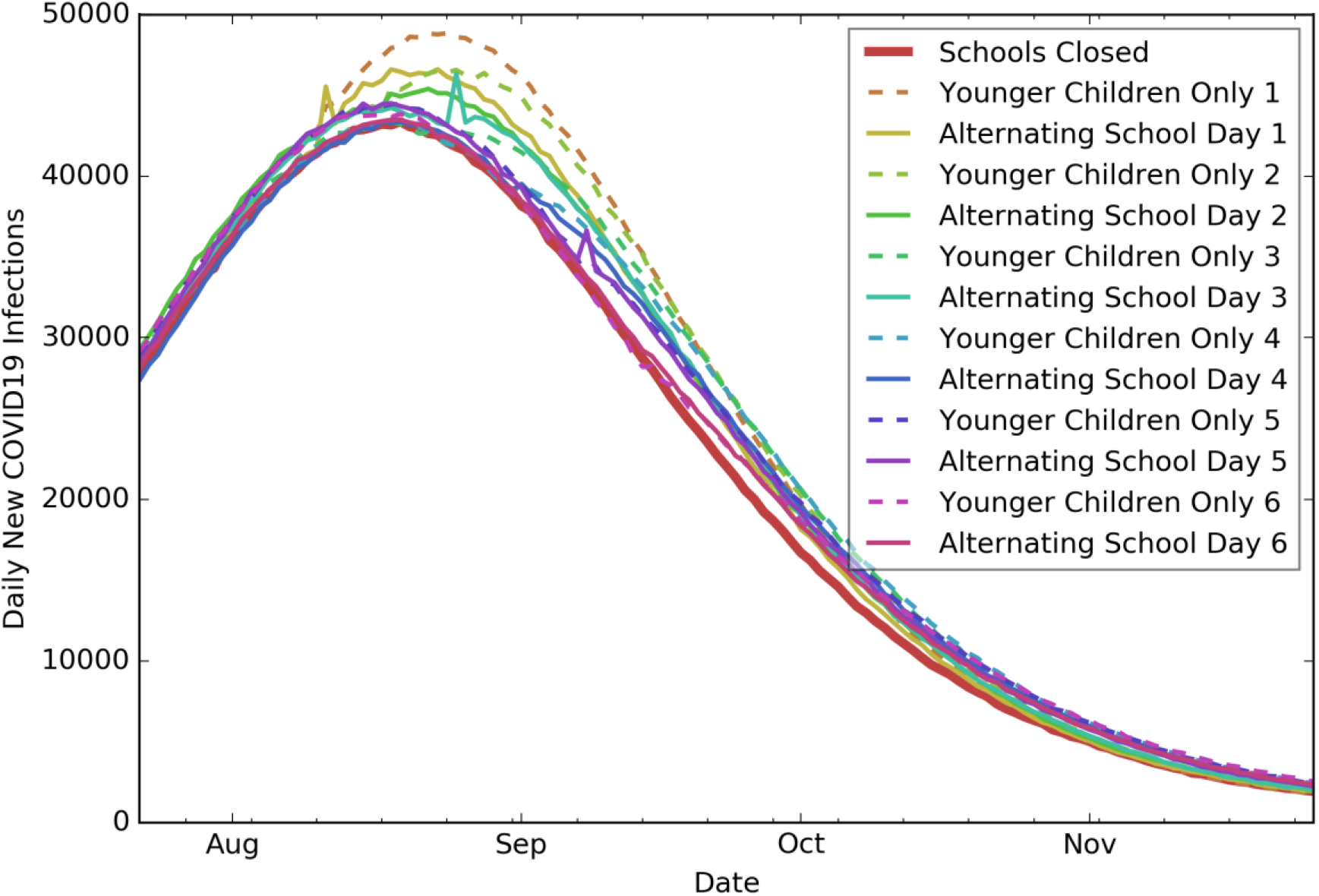
Daily new COVID19 infections under the *schools closed* schedule, the *younger children only* schedule, and the *alternating school day* schedule. The *alternating school day for younger children* schedule is similar to the *alternating school day* schedule.

## Discussion

Governments and school systems have been grappling with the decisions of how to prepare students for academic success while also trying to minimize the spread of COVID19. The negative impact of school closures has been disproportionately high on some students, e.g., those who do not have access to technology in the household, lack proper childcare, face an unsafe home environment, or have traditionally relied on the school system for meals, special education, counseling, and other forms of social or emotional support. While children seem to be less affected by COVID 19 than adults, they could be transmitters of COVID19, potentially increasing community infection spread if schools were to return to in-person instruction (16), particularly if the implementation of social distancing measures and recommendations remains financially or physically challenging for some schools (e.g., poor ventilation in buildings, short supply of disinfectant products, state budget shortfalls, etc.) (1).

There has been considerable debate about the benefits and risks of when and how to return to in-person instruction in schools during fall 2020. The American Academy of Pediatrics “strongly advocates that all policy considerations for the coming school year should start with a goal of having students physically present in school (18).” Some school systems delayed their opening dates or announced fully online instruction for the fall semester, while others considered hybrid models such as “groups of students to attend on alternating days or weeks, as well as allowing only younger students to attend while older students learn at home (19).”

According to our study results, delaying the reopening date would have a minimal impact on the peak day and peak number of new infections under the *alternating school day* schedule, the *younger children only* schedule, and the *alternating school day for younger children* schedule. However, under the *regular* schedule, delaying the reopening date from August 10 to September 17 could delay the peak day by 26 days and reduce the peak number of new infections from approximately 67.9 to 43.4 thousand.

The cumulative number of infections as well as the percentage of the population infected at the peak are similar under the *alternating school day* and the *schools closed schedules*, and significantly lower compared to the *regular schedule*. Hence, implementing an *alternating school day schedule* or limiting interactions between student cohorts during the in-person instruction could have a significant impact on slowing down the disease spread.

The *younger children only* schedule does not result in a significant reduction in cumulative infections compared to the *alternating school day* schedule. The *alternating school day for younger children* schedule reduces the number of infections compared to the *younger children only* schedule but not as much as the *schools closed* schedule.

COVID19 has had a significant impact on the society both in terms of public health and social and economic interactions. The health and well-being of the population are of the utmost importance, but there is also a growing desire to return to in-person instruction to support the educational development of children.

As school systems continue to develop plans for reopening, it is critical to understand the impact of various reopening scenarios on public health as well as the children’s development and the economy. Our results suggest that reopening schools following a *regular* schedule, i.e., all children returning to school without strict public health measures, would significantly increase the number of infections and deaths, i.e., have serious negative public health consequences. The *alternating school day* schedule, especially if offered as an option to families and teachers who prefer to opt in, provides a good balance in reducing the infection spread compared to the *regular* schedule, while ensuring access to in-person education.

This study did not consider the use of face masks or testing and tracing; if these measures were incorporated into the model, this would likely lead to a lower number of infections and deaths in all scenarios, due to the reduced transmission rate, but not change the relative ordering of the scenarios regarding infections and deaths. Regardless of how school instruction is formatted in the fall, it is important to continue promoting physical distancing measures and the usage of face masks as well as establishing testing and tracing practices to ensure prevention or early detection of outbreaks in schools.

## Materials and Methods

The study population consisted of children, youth, adults, and elderly stratified by age groups: 0-9, 10-19, 20-64, and 65+ in the state of Georgia.

The results were obtained by adapting and utilizing an agent-based simulation model to predict the spread of COVID19 geographically and over time (20–23). The model captured the progression of the disease in an individual and interactions within households, workplaces, schools, and communities. It enabled the testing of scenarios incorporating various types and durations of physical distancing interventions, namely school closures, shelter-in-place, and voluntary quarantine of households (i.e., the entire household remains home if there is a person in the household with cold/flu-like symptoms) as well as the public’s compliance levels. The model was populated using data from the Census Bureau (17, 24, 25) for the demographic and workflow information at the census tract level in Georgia and initialized with infection “seeds” following the distribution of the total number of confirmed COVID19 cases in Georgia (as of May 15^th^) at the county level (26) using the Huntington-Hill method of apportionment (27). Additional information about the model and the data sources can be found in (20).

On each day, the school status (“attending in-person or “attending online”) of younger children (0-9) and older children (10–19) was tracked and updated in the simulation depending on the reopening scenario. Children “attending online” did not engage in school-based peer interactions.

In the tables and figures that follow, scenarios were labeled by their names, as well as numbers 1 through 6 which refer to reopening schools on August 10, August 17, August 24, August 31, September 7, and September 14, respectively. For example, *Alternating School Day 3* refers to the scenario in which schools are reopened on August 24 and children adhere to an *alternating school day* schedule as defined in the Introduction section. All scenarios tested were built upon the base scenario, described in *Figure 1*.

The results presented were for the time period of February 18, 2020 to November 24, 2020. The simulation incorporated school closures during March 16-August 10 (28) and the following physical distancing practices with varying levels of compliance:

- Shelter-in-place: Staying home and refraining from interactions outside of the household. In Georgia, shelter-in-place order was in place during April 3-30, 2020. Shelter-in-place compliance of 80% was assumed for that time period.
- Voluntary quarantine: An entire household stays home if there is a person in the household with cold/flu-like symptoms, until the entire household is symptom-free. Voluntary quarantine compliance was 30% in mid-February, increased by 10% weekly until mid-March, and remained at 60% until the end of April. After the end of shelter-in-place, voluntary quarantine compliance was 70% and decreased by 10% weekly until stabilizing at 20%.
- Voluntary shelter-in-place: An entire household chooses to remain home, regardless of whether they have symptoms or not. During the week after the end of shelter-in-place, voluntary shelter-in-place compliance was 60% and decreased to 40%, 20%, and 5%, in consecutive weeks, until stabilizing at 5%.
- Voluntary shelter-in-place and voluntary quarantine compliance levels in the model were chosen to be in line with social mobility indicators (29).

The infection spread outcome measures over the time horizon of the study included:

- <u>Cumulative deaths</u>: cumulative number of people who died due to COVID19 over the time horizon of the study.
- <u>Cumulative infections</u>: cumulative number of people infected (including asymptomatic infections).
- <u>Infection attack rate (IAR)</u>: cumulative percentage of the population infected.
- <u>Peak day</u>: the day when the number of new infections was highest.
- <u>Peak infection</u>: the number of the population infected on the peak day.

## Data Availability

All data used in this study is publicly available.

https://dph.georgia.gov/covid-19-daily-status-report

## Acknowledgments

The research was supported by the William W. George and by the Virginia C. and Joseph C. Mello endowments at Georgia Tech. This research was supported in part by NSF grant MRI 1828187 and research cyberinfrastructure resources and services provided by the Partnership for an Advanced Computing Environment (PACE) at the Georgia Institute of Technology. The authors of this paper are thankful to state representatives for sharing multiple data sources for the confirmed cases in Georgia. The authors are also thankful to Georgia Tech students and colleagues Melody Shellman, Hannah Lin, Ethan Channel, Pravara Harati, April Yu Zhuoting, Gabriel Siewert, and Christopher Stone for their contributions and support.

## Supplementary Information for

### This PDF file includes

Legend for Dataset S1

### Dataset S1 (separate file)

Cumulative infections and deaths under all scenarios presented in this study.

